# Herpes simplex virus genomes from an under-sampled population in Namibia reveal novel genetic diversity

**DOI:** 10.64898/2026.02.18.26346525

**Authors:** Christopher D. Bowen, Alexandre Blake, Daniel W. Renner, M. Ashley Hazel, John Jakurama, Justy Matundu, Moriah L. Szpara, Nita Bharti

**Affiliations:** Department of Biology, The Center for Infectious Disease Dynamics, Huck Institutes for the Life Sciences, Pennsylvania State University, University Park, PA; Department of Biochemistry and Molecular Biology, Pennsylvania State University, University Park, PA; Institute for Health & Aging, School of Nursing, University of California, San Francisco, CA; Kaoko Information Centre, Opuwo, Namibia

## Abstract

Herpes simplex virus (HSV) is an endemic pathogen, infecting most adults world-wide. HSV infection can cause a wide spectrum of disease outcomes, ranging from asymptomatic infection or mild lesions to rare cases of infectious keratitis, encephalitis, and death. HSV genome sequences have been shown to differ between individual patients, as well as within individuals. To date, the vast majority of publicly available HSV genomic data has come from Europe and North America. Our current understanding of these patterns are missing data from under-sampled populations, particularly in South America, Africa, and Asia. Also missing have been HSV samples from non-industrial (e.g., agricultural, pastoral) populations, for which the natural environment plays a large role in health and disease dynamics. In this study, we capitalized on Whatman FTA card stabilization of DNA to develop a procedure for capturing oral and genital swabs from a geographically isolated pastoralist population in a desert region of northern Namibia. These are the first data to document HSV diversity in this type of remote setting. These are also the first HSV genomes from Namibia. These approaches may prove useful in broadening the accessibility of viral detection for these chronic pathogens, help improve diagnostics, and raise public health awareness about the burden of these pathogens in under-served populations.

## Introduction

Herpes simplex viruses (HSV) are large double-stranded DNA (dsDNA) viruses classified in the *Alphaherpesvirinae* sub-family of the *Herpesviridae* family [1]. There are two distantly-related species, HSV-1 and HSV-2 (*Simplexvirus humanalpha1 or 2*), that can infect the human oral or genital niches. HSV-1 has established lifelong infections in about 67% of adults worldwide, or an estimated 3.7 billion people under the age of 50, while HSV-2 has infected an estimated 491 million [2]. While primary infection by HSV occurs at the mucosal epithelium, lifelong latency is established in the nervous system [1]. From this neuronal reservoir, the virus can reactivate in response to a number of stressful stimuli. Reactivation enables the virus to return to the skin surface, initiate new shedding and/or lesions, and potentially spread to new hosts.

Previous studies have shown that HSV isolates from unrelated individuals can differ between 2-4% in DNA identity genome-wide [3,4]. HSV-2 appears to have a more recent evolutionary origin compared to HSV-1 and it has concomitantly lower diversity between isolates [4–6]. Both HSV species have a large dsDNA genome of over 150 kb, which encodes >75 proteins. This amount of variation can produce distinct coding differences in >60 proteins between any two unrelated isolates of HSV-1 [3,4,7]. HSV-1 has been shown to acquire genetic diversity by a variety of mechanisms, notably fluctuation in tandem repeat (TR) lengths, insertions/deletions (in/dels), single nucleotide polymorphisms (SNPs), and recombination events. Isolates from participants of similar geographic origins tend to group together in phylogenetic analyses, although these groupings show intermingling with the addition of recent isolates [3,4,6–15]. It is crucial to understand the amount of genetic diversity circulating worldwide to develop broadly effective diagnostics and treatments.

To date, the vast majority of HSV-1 isolates sequenced have been collected from urban clinics in either North America or Europe [3,7,11–15]. There is a significant underrepresentation of samples from South America, Africa, and Asia as well as from subsistence populations and rural areas globally [8–10,16,17]. It is necessary to sample from these regions and groups to characterize the entirety of genetic diversity present in circulating HSV-1. Geographically isolated populations are difficult to sample, and the long-term chronic nature of HSV infections may lead to tolerance of symptoms, dissuading individuals from seeking formal diagnoses or ongoing treatment, which increases the difficulty of sample collection.

In this study, we quantify HSV shedding in a pastoralist population in Namibia through two field studies conducted in 2015 and 2016. These data include individuals and families from the Kaokoveld region of northwestern Namibia. The area is a sparsely populated desert, with the vast majority of participants self-identifying as members of the Himba tribe. A small number of participants identified with additional tribes residing in the area, including the Tjimba, Ovambo, Zemba, and Twe [18–20] all of whom are Bantu-speaking with cultural ties to semi-nomadic pastoralism, supported by subsistence agriculture. In the typical gender-based division of labor in Kaokoveld, men primarily tend cattle herds, which requires frequent dispersal from home villages to find grazing and water. Women manage subsistence farms, goat herds, and childcare. Although men are more mobile than women, it is not uncommon for residents of this region to be away from their primary residence for many weeks, either for subsistence or to visit extended family. Partner concurrency is common and tolerated for men and women and occurs through extra-marital relationships and polygynous marriage.

This study was part of a larger study about community health, access to health care, contact networks, and movement in pastoralist regions of Namibia [18–21]. Prior studies in this population have already quantified the overall high seroprevalence of HSV-2 and incidence of other sexually transmitted infections, although there is no prior data on HSV-1 prevalence in this region [18–21]. We extended invitations to enroll in the study regardless of exposure to HSV or symptoms of HSV. We interviewed participants about their interpersonal contacts, access to healthcare, movement, and basic demographic information. Participants also provided self-collected swabs of oral and genital areas. These samples were later analyzed for HSV DNA detection. For samples with sufficient DNA, we completed viral genome sequencing and comparative genomic analysis. We paired viral DNA detection with contact-network analysis to integrate data on HSV shedding with human interaction networks. Although detectable HSV shedding was sparse in this study, these approaches demonstrate proof of principle that viral sampling and storage is feasible in remote settings and illustrate the potential to integrate data on human and viral interaction networks.

## Results

### Sampling from geographically isolated pastoralists in Namibia

Samples were collected in the Kaokoveld region of northern Namibia (**Figure 1A**). These efforts spanned two site visits, one in February 2015 that occurred during a long period of drought, and the second in October 2016 that followed a period of adequate rainfall. Participants were enrolled in the study regardless of HSV symptoms or exposure.

**Figure 1:**
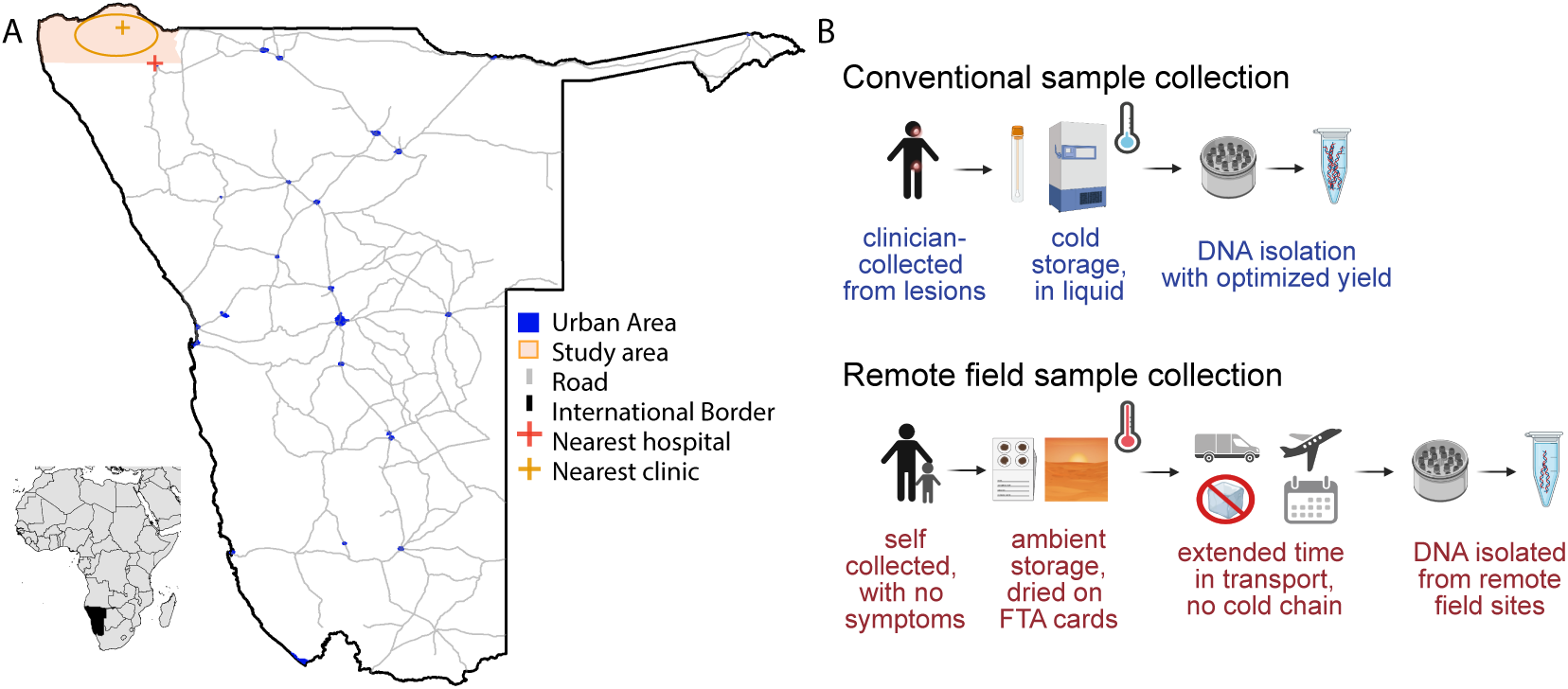
Maps of study area and collection of viral DNA. **(A)** Lower left inset: Namibia shaded black. Enlarged map: Urbanized areas in blue, approximate location of study areas in orange oval in the north. Location of nearest clinic shown with orange cross, nearest hospital shown with a red cross. Major roads in grey lines (from World Bank). **(B)** Top row: conventional clinical HSV sample collection from lesions; bottom row: remote field sample collection in the present study. Diagram components in B created in BioRender.

Study participation for adults entailed an interview and self-collection of oral and genital samples for detection of viral DNA. For each juvenile participant in both field studies, an adult guardian collected a single oral swab only and provided basic demographic and familial information (29 in 2015, 28 in 2016; details below). In 2015, we completed full length interviews with 75 adults, of whom 37 were male [M] and 38 female [F]) (we collected additional samples paired with demographic data only from adults, details below). Of these individuals, 1 M and 1 F reported experiencing genital lesions in the previous 12 months that may have been consistent with HSV infection, while 15 M and 8 F reported experiencing potential HSV oral lesions in their lifetime. In 2016, a total of 65 adults completed interviews and provided samples (31 M, 34 F). Of the 2016 study participants, 0 M and 3 F reported experiencing potential genital lesions in the previous 12 months, and 21 M and 5 F reported experiencing potential HSV oral lesions in their lifetime. Zero study participants reported lesions at the time of the swab collection. HSV DNA detected in this study likely reflects asymptomatic shedding.

In the absence of cold-storage, we developed a low-technology approach to store and transport viral samples for later HSV detection and DNA isolation (**Figure 1B**). Participants swabbed their oral and genital areas, and the swabs were immediately saturated in preservative-free saline and applied to Whatman FTA cards [22]. These FTA cards trap nucleic acids present in the sample, inactivating potential pathogens. They are ideal for safe handling, ambient storage, and compact, light-weight transport. During both the 2015 and 2016 field studies, adult participants self-collected samples from their external oral and genital areas (see Methods for details, and **Figure 2** for overview of samples). In 2016, we added the self-collection of internal oral (mouth) swabs for males and females, and internal genital swabs for adult females. A total of 486 samples were collected, with n=229 from the 2015 field season, and n=257 from the 2016 field season (**Figure 2**).

**Figure 2.**
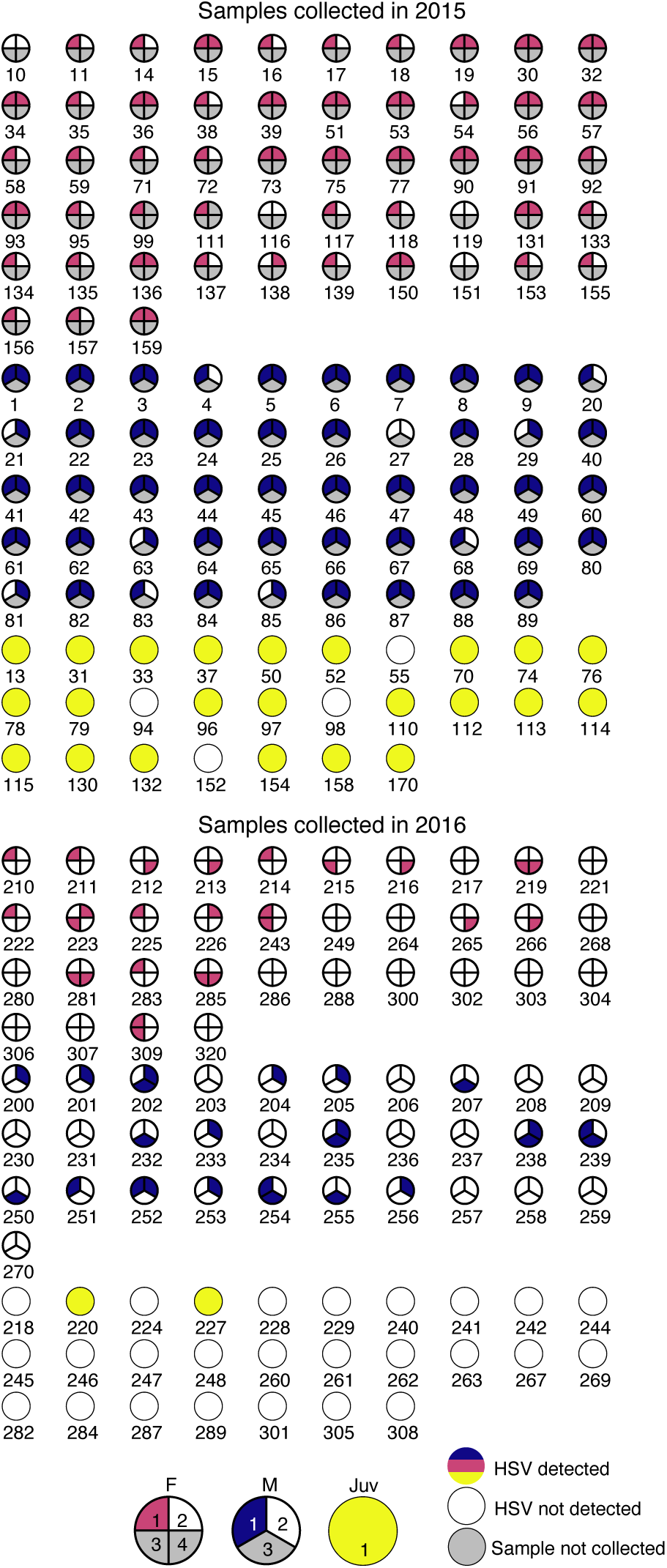
Plots of self-sampling for all participants during 2015 (above) and 2016 (below) data collection efforts. Samples were numbered from 1 to 4 as follows: 1 = oral external; 2 = genital external; 3 = oral internal; 4 = genital internal. Samples 1 and 2 were collected from all adults in all years, sample 3 was collected in 2016 only from all adults, sample 4 was collected in 2016 only from all adult female participants. A maximum of two oral samples (1, oral external, and 3, oral internal) and two genital (2, genital external, and 4, genital internal) samples were taken from female adults, and their plots are divided into corresponding quarters. A maximum of two oral samples (1, oral external, and 3, oral internal) and one genital sample (2, genital external) were taken from male adults, and their plots are divided into corresponding thirds. Only one sample was taken from juveniles (1, oral cheek swab). If HSV was not detected in the sample, the corresponding part of the plot is white, and if HSV was detected, the corresponding part of the plot is red for adult females, blue for adult males, and yellow for juveniles.

We used quantitative PCR (qPCR) for HSV DNA to explore the potential for detection of asymptomatic viral shedding. After DNA isolation from the Whatman FTA cards, we quantified the amount of total DNA and the copy number of HSV genomes in each sample (by Qubit and qPCR respectively; see Methods for details). A total of 233 of the 486 samples analyzed (48%) had detectable HSV DNA. From these, we detected >1,000 viral genomes in only 22 samples (4.5%) (**Table 1**). The total sample pool (n=229) collected in 2015 included 98 samples from 49 males; 102 samples from 51 female participants; and 29 samples from 29 juvenile participants (**Figure 2**). Of these samples collected in 2015, we detected >1,000 viral genomes in 18 samples (**Figure 3A,C**). From the 2016 field season, the sample pool (n=257) included 93 samples from 31 male participants; 136 samples from 34 female participants; and 28 samples from 28 juvenile participants (**Figure 2**). Of these samples collected in 2016, we detected >1,000 viral genomes in just 4 samples (**Figure 3B,D**). There was no apparent correlation between the total amount of DNA and the number of HSV genomes detected in each sample (**Figure 3**).

**Figure 3:**
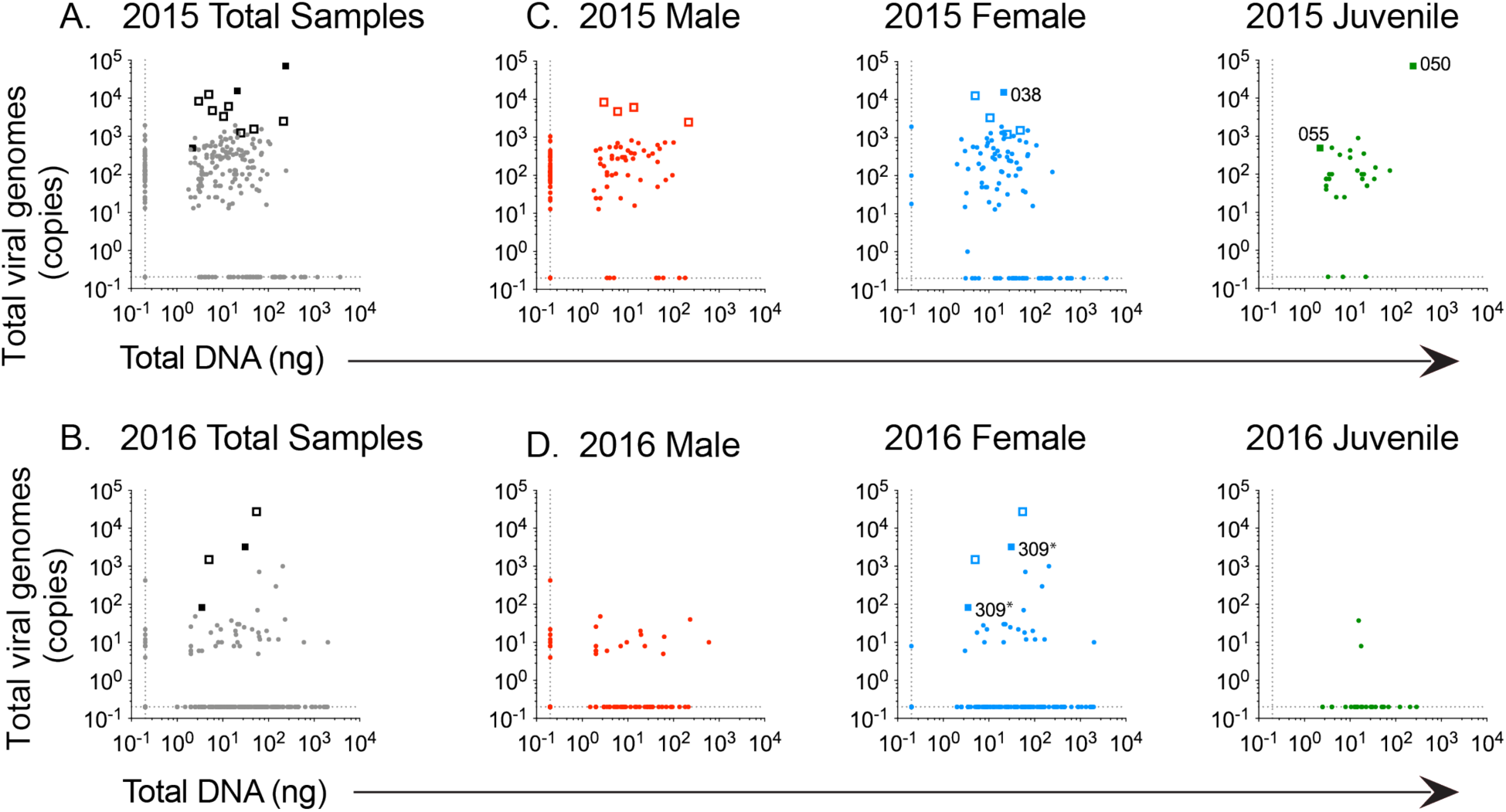
Quantification of total DNA and viral-specific DNA extracted from participant samples. Total DNA (in ng) isolated from FTA cards (x-axis) was compared to the total amount of virus-specific DNA (y-axis; viral genome copy number). Total DNA was quantified by fluorimetry and HSV-specific DNA was quantified by qPCR (see Methods for details). Data are shown for samples from the **(A)** 2015 field study and **(B)** 2016 field study. Samples for which viral genome sequencing was attempted are denoted as hollow boxes. Solid bold boxes indicate successfully sequenced samples. Source labels for the successfully sequenced viral samples are indicated on specific points in **(C,D)**. Subsets of these data are plotted separately for **(C,D)** male, female, and juvenile samples for each field study year. The 2015 field study included 229 samples: 98 swabs from 49 males; 102 swabs from 51 females; and 29 swabs from juveniles. The 2016 field study included 257 samples: 93 swabs from 31 males; 136 swabs from 34 females; and 28 samples from juveniles. Two samples (oral-external and genital-external) from study participant 309 were pooled to obtain sufficient viral DNA for oligo-enrichment and sequencing.

**Table 1:** Total DNA yield and number of HSV genomes detected in remote field samples.

| Samples | N.D.<br>(<1 ng DNA) | 1-100 ng<br>DNA | >100 ng<br>DNA | N.D.<br>(<1 viral<br>genome) | 1-1,000 viral<br>genomes | >1,000 viral<br>genomes |
| --- | --- | --- | --- | --- | --- | --- |
| **Total (2015)**<br>**n= 229** | 41 | 167 | 21 | 47 | 164 | 18 |
| **Total (2016)**<br>**n= 257** | 49 | 164 | 44 | 206 | 47 | 4 |
| **Total (All)**<br>**n= 486** | 90 | 331 | 65 | 253 | 211 | 22 |

### Sequencing of samples with high HSV genome copy numbers

We selected samples with the highest HSV copy number (**Table 1**) for our attempts to sequence viral genomes from remote field samples. Sequencing libraries were constructed as previously described [23], and samples were subjected to Illumina paired-end deep sequencing. HSV genomes were assembled using an in-house *de novo* viral genome assembly workflow [24]. We successfully recovered viral genome from four samples (**Table 2**). Other samples attempted for viral genome sequencing yielded insufficient viral sequencing reads for viral genome assembly (**Figure 3**). The four HSV-1 viral genomes were all derived from oral samples (**Table 2**). The average coverage depth ranged from >12,000× (sample 038) to 39× (sample 309). Three of these viral genomes (038, 055, and 050) had >100X coverage across a majority of the genome: HSV1-Namibia-2015-oral038 (12,866× coverage), HSV1-Namibia-2015-oral055 (722×), and HSV1-Namibia-2015-oral050 (142×). For participant 309, the sequencing libraries of separate oral-external and genital-external libraries both yielded insufficient viral reads to reconstruct independent viral genomes. Instead, we pooled viral sequencing reads across these two samples (23.5k total), which was sufficient to recover a low-coverage (39×) viral genome from this participant: HSV1-Namibia-2016-309combined. To our knowledge, these four viral genomes represent the first HSV-1 genomes derived from Namibia.

**Table 2:** Data and sequencing statistics for samples with successful HSV-1 genome recovery.

| Sample<br># | Sample source | Field<br>season | Average<br>coverage<br>depth | Mapped<br>reads | %<br>genome<br>coverage<br>>100× | GenBank<br>Accession<br># |
| --- | --- | --- | --- | --- | --- | --- |
| 038 | Oral external, adult female | 2015 | 12,866× | 7.0 M | 99% | PP379031 |
| 055 | Oral external, juvenile | 2015 | 722× | 8.8 M | 97% | PP379033 |
| 050 | Oral external, juvenile | 2015 | 142× | 83.5 K | 65% | PP379032 |
| 309* | Oral external/genital<br>external, adult female | 2016 | 39× | 23.5 K | 3% | PP379034 |

\* DNA was pooled for two HSV-positive samples (oral-external and genital-external) from study participant 309, to provide sufficient viral DNA templates for oligo-enrichment and sequencing.

### Comparative genomic analysis of Namibian HSV-1 isolates

To place these new viral genomes from Namibia into a global context, we compared their relatedness to previously described HSV-1 genomes from Africa and from all available geographic regions. There are currently only 16 HSV-1 strains from the entire African continent. Fifteen of these were collected in Nairobi, Kenya between 1981-1984 [8] and sequenced almost 30 years later [3]. Additionally, a South African HSV-1 genome collected from a 2008 blood sample was deposited in GenBank [25].

We first aligned and compared the four uncultured Namibian HSV-1 isolates to these 16 strains from elsewhere in Africa (**Figure 4A**). We then extended this comparison to a globally representative set of 66 strains (**Figure 4B**; see **Supplemental Table 1** for list of strains). In both network graph analyses, the four Namibian strains fit within the scope of viral genetic diversity previously defined by the African strains, which have been noted in prior studies to already encompass a high level of all known HSV-1 genetic diversity [9,10]. The HSV-1 genomes from participants 050 and 055 clustered closely on both network graphs. We also compared the protein-coding and intergenic regions of these four Namibian HSV-1 isolates vs. the set of 66 globally distributed strains. This comparison revealed no variations shared among all four Namibian HSV-1 isolates that distinguished these isolates from others. Rather all coding and non-coding variations in these genomes reflected constellations of those observed in prior African and globally distributed genomes.

**Figure 4:**
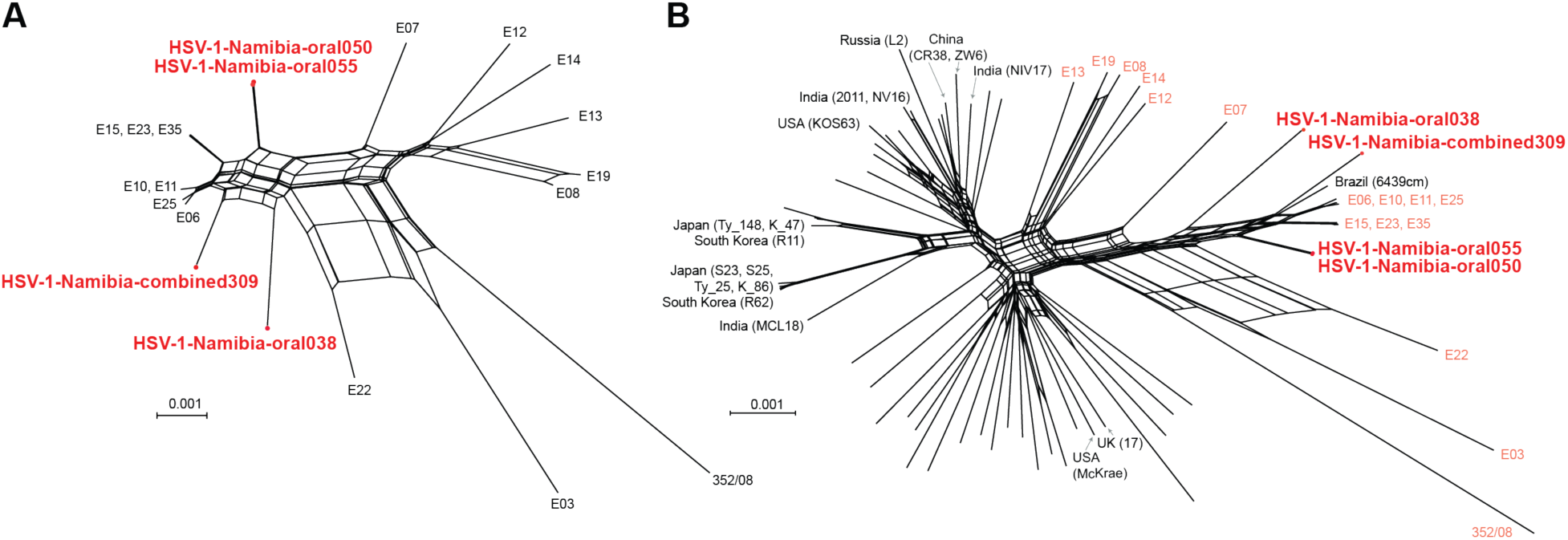
Network graph analysis illustrates the relatedness of new Namibian field isolates to previously sequenced African and other HSV-1 isolates. The four new Namibia HSV-1 genomes (red) were aligned and compared in **(A)** a SplitsTree network graph analysis with other African strains (**B**) or a globally representative set of all known HSV-1 genomes. All but one of the African strains were collected in Nairobi, Kenya between 1981-1984, as part of a study by Sakaoka et al [8]. One sample, 352/08, was collected in 2008 in South Africa [25]. In the global network (**B**), other HSV-1 genomes from Africa are highlighted (pink). Network tips are labeled for viral genomes of Asian origin and for representative strains from the USA (KOS63, McKrae) or Europe (17). The inner reticulations on the network graph indicate signs of past recombination between strains. **Supplemental Table 1** lists the previously sequenced HSV-1 strains used for this comparison.

### Integrating HSV detection with human contact networks

Study participants completed interviews at the time of the collection of samples for HSV-detection, providing information on socio-demographics, access to health care, movement patterns, and close interpersonal contacts within and between the settlements in the region [26]. Survey participants self-reported social and sexual contacts for the six months preceding the sample collection. In the resulting contact network (see Methods for details), the study participants and their named sexual contacts comprise the nodes and the edges represent contacts between nodes, including family and sexual-partner interactions (**Figure 5**). The network of participants was strongly connected; the majority of the edges linked sexual contacts (light grey edges on **Figure 5**); 82.3% (153/186) and 75.6% (195/254) in 2015 and 2016, respectively. The family/household ties among the participants (dark grey edges on **Figure 5**) made up the remainder of the edges.

**Figure 5:**
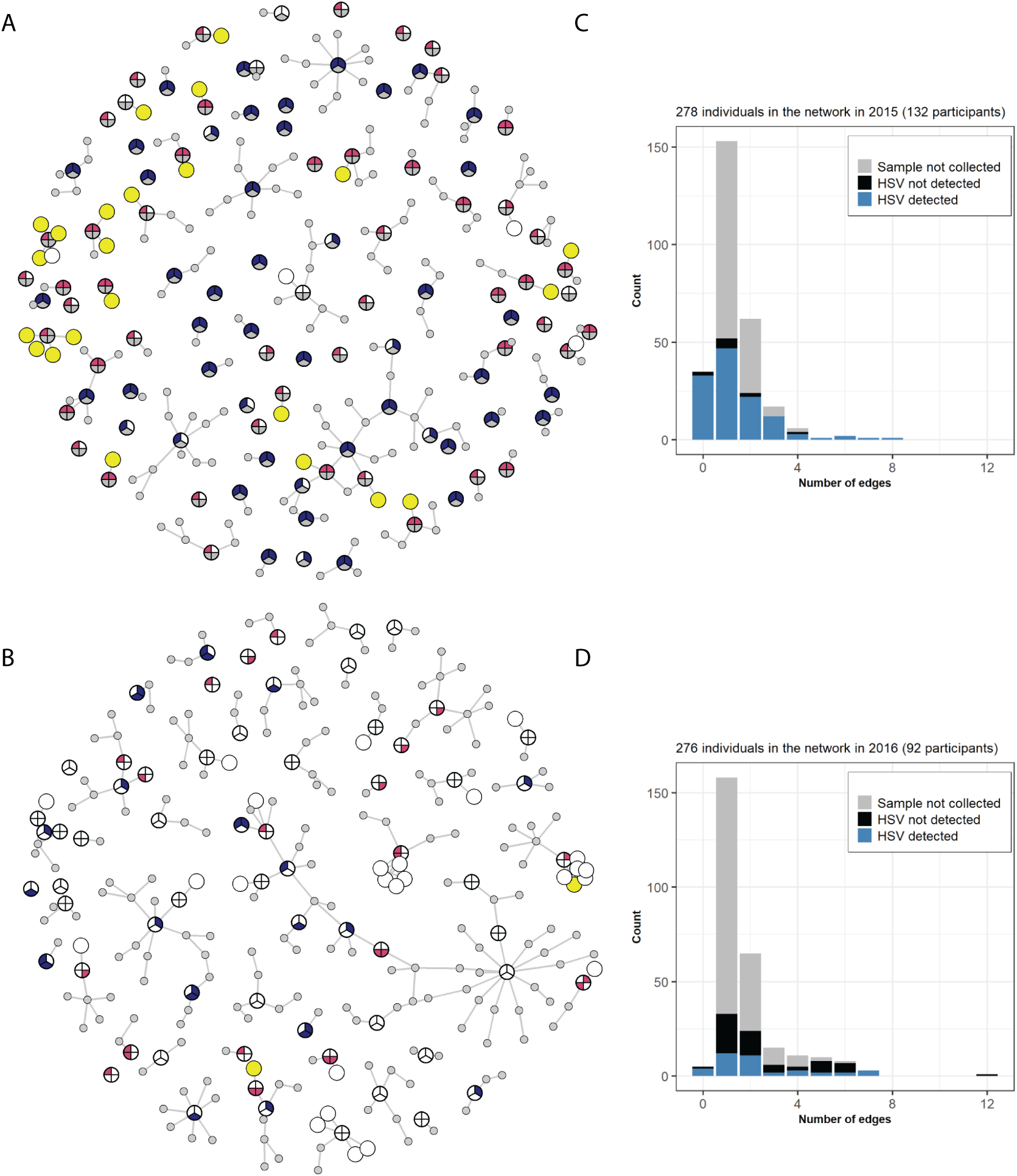
Contact network of study participants. Edges show family ties or sexual contacts for participants sampled in **(A)** 2015 and **(B)** 2016. The grey nodes represent named sexual contacts who were not sampled for HSV sequencing and did not participate in the survey. See Figure 2 for full details of circular sampling-diagrams. If a participant was not sampled at a particular site, the corresponding part of the node is shaded in grey. If HSV was not detected in the sample, the corresponding part of the node is white, and if HSV was detected, the corresponding part of the node is red for female adults, blue for male adults, and yellow for juveniles. The inset bar plots for **(C)** 2015 **(D)** and 2016 show the distribution of the number of edges (family ties or sexual contacts) broken down by HSV detection (light blue = HSV detected, dark blue = not detected, grey = not sampled).

We completed a descriptive analysis of the networks focusing on degree distribution, shortest paths, and betweenness centrality. The degree measures the number of individuals to which each individual is linked, or the potential connections along which a transmissible pathogen could spread. We expected shorter paths between infected participants and that infected participants would have a higher betweenness centrality. The average degree distributions in 2015 and 2016 were 1.35 and 1.84, ranging from 0 to 8 and 0 to 12. The network highlights that sharing sexual partners was not uncommon. We identified sexual partners who participated in the study and were named by one or more study participants. Across both years, the distribution of the length of the shortest paths between participants with HSV shedding detected did not differ significantly from the length of the shortest paths between the participants from whom HSV shedding was not detected (p=0.9 and p=0.4) (**Figure 4**, inset graphs). Similarly, participants with detected HSV shedding did not have a significantly higher betweenness centrality compared to the participants without shedding (p=0.8 and p=0.5). However due to the nature of our HSV detection being limited to a single time point and only asymptomatic shedding at best, we anticipate that these data under-report the true number of HSV-positive individuals in the population [19]. Future studies that combine these data with seroprevalence detection of past infection may reveal different patterns of degree distribution and betweenness centrality.

## Discussion

In this study, we examined HSV shedding in a pastoralist population in the sparsely populated desert region of Kaokoveld, in northwestern Namibia. There are no prior statistics on HSV-1 shedding in this population; these data represent the first measures of this public health burden of this pathogen. Prior studies of sexually transmitted infections (STIs) in this region found a high seroprevalence of HSV-2 and frequent PCR-detection of the bacterial pathogen gonorrhea [18,19,21]. While some study participants self-reported either oral or genital lesions compatible with HSV infection in the year preceding sample collection, none reported active lesions at the time of sample collection. Using self-collected swabs immobilized on FTA cards, we successfully isolated viral DNA from oral and genital swabs, and we successfully recovered viral genomes from four of these samples.

The low technology approach of using FTA card immobilization allowed us to screen a previously under-sampled and hard to reach population (**Figure 1**) and successfully recover HSV genomes from remote locations. Across two years of sample collection, we found that 48% of samples (233 total) had detectable viral DNA present, but just 4.5% (22 total samples) had 1,000 or more viral genomes detected. This level of viral shedding and detection is not surprising, given a single time-point of sample collection and lack of symptomatic lesions reported by these participants. The low levels of HSV DNA detected in this study reflect the findings of prior studies on asymptomatic HSV shedding [27–29]. However this level of viral shedding represents the lower threshold of viral genetic material needed for successful genome sequencing, even when using oligonucleotide-bait based enrichment methods [13,14,30,23]. This physical limitation of the starting material likely constrained our recovery capability, resulting in only four viral genomes from 233 samples with detectable viral DNA. Future studies targeting sample collection from symptomatic participants would likely yield a greater number of HSV-positive samples with higher levels of HSV DNA per sample, enabling higher rates of viral genome recovery. However, this study demonstrates that samples collected from isolated populations in harsh environmental conditions can be successfully recovered and sequenced, providing new avenues to address the current disparity in our knowledge of HSV genetic diversity from under-sampled regions of the world, and from remote or rural populations globally.

We collected samples from the same two settlements across two years, however these populations are highly mobile and these data represent individuals and families from many locations and settlements across Kaokoveld. The area had been experiencing severe drought during our sampling efforts in 2015, leading to massive cattle die offs and limited food availability. Although our sampling efforts in 2016 occurred during the dry season, there had been sufficient rainfall prior to our arrival, which reduced ongoing herd die-off. More goats were present than the more prized, but less drought-resistant, cattle, and subsistence maize gardens had been more productive than in 2015. Across samples collected during the drought-phase study in 2015, we detected any HSV (>1 genome) in 81% of individuals, but only 20% for 2016. Likewise, there were 18 samples with >1,000 HSV genome copies in the 2015 field season, but only 4 such samples in 2016 (**Figure 3**). It is unclear how these environmental stressors may impact HSV shedding. One hypothesis is that drought-induced social and health stress lead to higher rates of viral reactivation and shedding, which would be consistent with our observations from 2015 and 2016.

Our goal for this study was to add a viral genetic component to a larger, ongoing investigation of the relationships between human movement, access to health care, health vulnerability, and data biases [18–21,26]. We studied remote pastoralist populations in Namibia to address these issues for multiple reasons. First, the location is difficult to access and has limited points of health care. As a result, these populations are largely overlooked in national health population and data from Namibia and globally [31]. Second, the cultural acceptance of extramarital partnerships and polygynous marriage results in high levels of sexual partner concurrency (**Figure 5**) and transparency in partnerships. We have observed a high prevalence of STIs in this population, including high rates of HSV-2 and gonorrhea, as noted above [18–21]. Finally, a lack of prior data from this area on the prevalence of HSV-1, a common virus, and viral genetic data, highlighted the underrepresentation of these populations in such studies.

If HSV genomes had been recoverable from multiple individuals within the interconnected social and sexual networks of the study participants (**Figure 5**), it may have been possible to detect past transmission events by measuring current viral relatedness. However, the infrequent and low-level of viral shedding in these asymptomatic participants did not yield sufficient viral genomes to enable this analysis. Future longitudinal studies, involving both symptomatic sample collection and socio-demographic interviews as done here, may provide these insights into human and viral interaction networks. Such studies on STI prevalence and its impacts on rural populations would also enable better public health interventions and help illuminate the interactions between environmental and microbial impacts on health.

## Materials and Methods

### Sample acquisition

The study design has approval from Penn State’s Institutional Review Board (IRB #STUDY00001510: Movement and Pathogens in Namibia) and Institutional Biosafety Committee (IBC #48898). We conducted surveys and biological sampling in two settlements in the desert of Kaokoveld, Namibia in 2015 and 2016. Upon arrival in the two Himba settlements, researchers sought and received approval from local settlement authorities at each location to explain the study. We gained their permission and assistance in recruiting volunteers. The sites of data collection were in the Kunene region, east of the Skeleton Coast and northwest of Etosha National Park, just south of the Angolan border. To access these locations, researchers arrived in Windhoek, drove approximately 8 hours to the small town of Opuwo, and then drove approximately 5 hours in a 4x4 vehicle on sand trails to arrive at the settlements where the study was done. Samples exit the study sites with the researchers. The lack of electricity and running water created a need for low-tech sample storage and transport capabilities. All personnel, research equipment, camping necessities, food, and petrol traveled in one vehicle, creating space and technology constraints for sample storage and transport.

Each survey and sampling session included exactly one researcher (either Bharti or Hazel) and one voluntary study participant as well as one translator, who read, wrote, spoke, and understood English and spoke and understood the Bantu language Otjiherero, the local language that all survey participants spoke. The translator was always the same sex as the survey participant; the female translator remained the same across both years, but each male translator held the role for exactly one field season. Study participants were restricted to adults. The designation of “adult” was locally determined by household responsibilities, interpersonal relationships, or age when known. In these settlements, individuals are considered adults at approximately 16 years of age. See below for detailed inclusion criteria.

We conducted surveys and collected samples with 102 adults during the rainy season in February 2015 across two Himba settlements. Of these 102 surveys, 27 surveys were part of a pilot phase during which only demographic data were collected with biological samples and full surveys were not conducted. We conducted 64 complete surveys paired with biological sample collection with adults during the dry season in October 2016 across the same two settlements. Participants were sampled from the same two Himba settlements each year. Some participants were residents in these settlements and others were visiting when the studies were conducted. Participants were specifically asked if they could recall when they most recently experienced typical symptoms of HSV 1 or 2 infection (oral or genital sores or lesions), if ever.

### Participant survey

Researchers Bharti and Hazel developed the survey instrument. Interviewers Jakurama and Matundu translated all items from English to Otjiherero, evaluated them for clarity and cultural context, and translated them from Otjiherero back to English. Bharti and Hazel reviewed the back-translations for clarity and made adjustments with Jakurama and Matundu as necessary to ensure consistent interpretations. Each interview began with an explanation of the survey process and purpose of the research, as well as opportunities for each potential participant to ask questions, decide whether to give their verbal consent for study participation, and to decline participation and revoke consent at any point. Enrolled participants were encouraged to ask questions for clarification, were permitted to skip or decline to answer any questions and could decline continuation at any point during the survey.

### Collection of biological samples on FTA cards

Along with each survey conducted with an adult, participants self-swabbed 2-4 tissue sites for this study. Each participant provided verbal consent for swab collection. Participants were given careful instructions, from a same-sex translator, and privacy behind a curtain, to self-collect swabs of the external genital area (2015 and 2016), and internal genital area (2016, female participants only). They also self-collected swabs of the external oral area including lips and saliva (2015, 2016), and internal oral area of the dorsal tongue (2016). Two drops of saline solution without preservatives were added to each swab after sample collection, which was then rolled onto a Whatman® Flinders Technology Associates (FTA^TM^) card for sample preservation and storage. The extent where the sample wetted the card was outlined in pencil, with a 2 mm buffer, while the sample was visibly wet. After the spots dried, cards were closed and stored in sealed envelopes with desiccant packs at ambient temperature (consistently over 40° C during the daytime). After the samples were placed on FTA cards, the swabs themselves were then discarded as biohazardous material. The study used sterile Copan FLOQSwabs®. Unique numeric codes were written on the FTA cards and the paper copies of the surveys so they could be paired later for analyses. FTA cards were returned to lab facilities at The Pennsylvania State University for DNA extraction, sequencing, and analysis.

### Study inclusion

Inclusion Criteria for participants were as follows: (1) All full participants must be culturally recognized adults who are sexually active. Thus, we recruited participants who were at least 16 years old. We did not have an age maximum for recruitment. (2) The study recruited women in equal proportions to men. Pregnant women were included, as this study posed no risk to them or to the fetus. (3) The population we worked with was ethnically homogenous. All the tribes who live in the areas where data were collected are ethnically, linguistically, or culturally related. (4) Visitors to the area from outside the settlements being studied and who were likely to be of different ethnic backgrounds were eligible for recruitment and were not excluded. (5) Verbal consent was obtained for all participants. With verbal consent from a guardian and from the juvenile, juveniles could complete a cheek swab and their guardian was asked for the names of their immediate family members (parents, siblings). Juveniles were not interviewed and did not provide a genital swab.

### DNA isolation from FTA cards

The area of the card where the sample was located (within the pencil outline) was cut out of the card with a razor blade. Total DNA was extracted from FTA cards using an adapted version of the manufacturer’s protocol [32]. Briefly, FTA card pieces were submerged in extraction buffer (10mM Tris-HCl, 10mM EDTA, 10mM NaCl, 2% Sodium dodecyl sulfate in water) and Proteinase K (Fisher). Samples were incubated overnight at 56° C with agitation. Total DNA was then extracted through multiple rounds of phenol-chloroform extraction, followed by ethanol precipitation. DNA pellets were dried via vacuum centrifuge and re-suspended in water. Total DNA was quantified by Qubit® 2.0 Fluorometer (Thermo Fisher). The amount of viral DNA was determined by a previously described quantitative real time PCR (qPCR) assay which amplified a region of the UL27 gene (which encodes glycoprotein B, or gB) and was quantified fluorescently [33,34]. This assay detects the UL27 of both HSV-1 and HSV-2, and it does not distinguish between the two. This quantification served as a measurement of total viral DNA present in each sample. Samples were considered “positive” for HSV if they had detectable viral DNA via qPCR (>1 genome, as compared to a standard curve of cultured HSV-1 DNA).

### Next-generation sequencing

Total DNA for each sample to be sequenced was sheared using a Covaris M220 focused-ultrasonicator (Covaris) under the following conditions: 10% duty, power 60, 200 cycles/burst for 60s at 4°C. Total DNA libraries were then prepared according to manufacturer’s protocols for the KAPA Biosystems HyperPrep Library Kit (with 14 cycles of amplification). Custom HSV-specific oligonucleotide probes [35] were used for in-solution target enrichment, using the Arbor Biosciences myBaits Target Capture Kit according to the manufacturer’s protocols [23]. The oligo-enriched library was amplified for 14 cycles using the KAPA HiFi HotStart Library Amplification Kit.

Samples were quantified by Qubit, HSV qPCR, and an adapter-specific qPCR, before multiplexing up to four libraries per flow cell. Libraries were sequenced using an Illumina MiSeq 600-cycle kit with v3 chemistry, according to the manufacturer’s recommendations. Prior to genome assembly, a BLAST database consisting of all known HSV-1 and HSV-2 genomes was constructed. This database was used for “positive selection” of HSV-like sequence reads (e-value < 10^-2^). These reads then *de novo* assembled using a published viral genome assembly (VirGA) pipeline for each sample, as previously described [24]. Briefly, after quality control, eight SSAKE *de novo* assemblies were generated and combined into a consensus genome using Celera, GapFiller, and mafnet [36]. Annotations were transferred from the HSV-1 reference genome (strain 17; GenBank accession JN555585) based on homology. GenBank accessions for these 4 genomes are listed in Table 1 ([PP379031-PP379034](https://www.ncbi.nlm.nih.gov/nuccore/PP379031,PP379032,PP379033,PP379034)). Raw sequence data are available via the Sequence Read Archive (SRA) under BioProject PRJNA1347855 (SRA accessions SAMN52875394-SAMN52875397).

### Viral genome network graph analysis

The four new Namibian HSV-1 genomes were aligned with other HSV-1 genomes using ClustalW2 [37]. **Supplemental Table 1** contains the full list of previously-published HSV-1 genomes used for these comparisons, along with their geographic source, GenBank accession, and reference(s). Multiple Alignment using Fast Fourier Transform (MAFFT) was used to construct pairwise global nucleotide alignments between trimmed genome sequences (i.e., terminal repeats were removed to avoid duplicated influence on the network graph). The Africa-specific alignment contains 132,024 ungapped bases (139,031 total in alignment), while the global alignment contains 122,753 ungapped bases (163,982 total in alignment). The MAFFT alignments were used to generate NeighborNet network graphs in SplitsTree4, using the uncorrected P distance and excluding gaps [38].

### Contact network among study participants

Using the information provided during socio-demographic surveys, we constructed a contact network for each year of data collection that encompassed household members, family members, and sexual contacts of the study participants. The socio-demographic survey information on family and sexual partners provided links between participants. In the contact-network (**Figure 4**), the study participants and their named sexual contacts comprise the nodes, and the edges are the reported contacts between nodes. Participants’ named sexual partners in the three months preceding data collection appear in the network as additional vertices and help provide a more complete picture of the connectivity in the study population. Named contacts who did not complete the socio-demographic interview were also not screened for HSV shedding. We used detectable HSV DNA (> 1 genome by qPCR) as an indicator of HSV infection for the contact-network analysis.

## Data Availability

All sequences have been deposited on GenBank and accession numbers provided in Table 2. All code for network analysis shown in Figure 5 will be available at https://github.com/bhartilab/namibia_hsv. Individual level data cannot be shared publicly (IRB #STUDY00001510). Aggregated data are available from Blake A, Hazel A, Jakurama J, Matundu J, Bharti N (2023) Disparities in mobile phone ownership reflect inequities in access to healthcare. PLOS Digital Health 2(7): e0000270. https://doi.org/10.1371/journal.pdig.0000270

## Acknowledgements

The authors obtained a research visa from the Namibian Ministry of Health and Social Services (MOHSS) and local institutional support through The University Center for Studies in Namibia (TUCSIN). Katuritara Tjiningire provided valuable translation assistance during 2016 as well as logistical support and community relationship building. Local authorities and all study participants provided valuable information, assistance, and samples, we are indebted to them for their help and participation. We thank members of the Szpara and Bharti labs for their support and feedback on this study and manuscript.

## Funding

The study was funded by The Human Health and the Environment Seed Grant Program from The Huck Institutes of the Life Sciences at The Pennsylvania State University and the Data to Insight/Data to Innovation Initiative (https://www.huck.psu.edu). NB acknowledges support from The Huck Institutes of Life Sciences early career professorship chair, The Branco Weiss Society in Science Fellowship (https://brancoweissfellowship.org), the joint National Institutes of Health (NIH) - National Science Foundation (NSF) - National Institute of Food and Agriculture (NIFA) Ecology and Evolution of Infectious Disease (award R01TW012434) (https://www.nsf.gov), and NSF RAPID (award 2202872)(https://www.nsf.gov). M.L.S., C.D.B., and D.W.R. acknowledge support of NIH R01 AI132692, R21 AI130676, and R01 AI163217 (https://grants.nih.gov). AH acknowledges support from the Wenner-Gren Foundation (https://wennergren.org). The funders played no role in the study design, data collection and analysis, decision to publish, or preparation of the manuscript.

## Notes

### Competing Interest Statement

The authors have declared no competing interest.

### Funding Statement

Yes

### Author Declarations

The study design has approval from Penn State's Institutional Review Board (IRB #STUDY00001510: Movement and Pathogens in Namibia) and Institutional Biosafety Committee (IBC #48898). Verbal consent was obtained from each participant.

